# Novel 3D imaging technology, as adjunct to mammography, improves Specificity markedly without reducing Sensitivity in BIRADS-4 patients

**DOI:** 10.1101/2025.11.27.25341183

**Authors:** V Marmarelis, N Michalopoulos, D Koulocheri, M Alafaki, M Sofras, E Groumpas, M Orme, G Zografos

**Affiliations:** Department of Biomedical Engineering, University of Southern California, Los Angeles, California, USA; Breast Unit of Hippocration Hospital, University of Athens School of Medicine, Greece; Mastoscopia SA, Athens, Greece & MUST Imaging Inc, Los Angeles, California, USA

**Keywords:** Breast cancer detection, Automated breast lesion differentiation, Non-ionizing 3D breast imaging, Operator-independent 3D breast imaging, Density-independent 3D breast imaging, BI-RADS 4 patients

## Abstract

**Objective:** To evaluate the potential diagnostic improvement accrued from using the novel 3D breast imaging technology of Multimodal UltraSonic Tomography (MUST) as adjunct to digital mammography for BI-RADS 4 patients referred to biopsy (clinical trial # EUDAMED/CIV-ID CIV-GR-22-05-039513).

**Methods:** MUST generates 3D tomographic images of pendant breast in water-bath using transmission-mode ultrasound measurements of acoustic refractivity and frequency-dependent attenuation. These measurements are fused via a properly developed algorithm into “advisory diagnostic images” (ADI) depicting the likelihood of malignancy at each voxel of the entire breast volume. In this clinical trial, MUST imaging was performed prior to biopsy on 207 BI-RADS 4 patients presenting micro-calcifications in mammography. The findings of the MUST ADI in the biopsy region were evaluated against the biopsy results

**Results:** Biopsy histopathology identified malignant lesions in 54 patients (26.2%). MUST ADI detected correctly all these malignancies (down to 2 mm in maximum dimension). In 31 of the 153 participants with negative biopsy (20.3%), MUST ADI found some “likely malignant” lesions within 20-mm radius from the putative point of biopsy. Breast density varied across the cohort, with 65.2% having dense breasts (ACR score 3-4). No dependence of MUST diagnostic performance on breast density was found in this cohort.

**Conclusion:** MUST imaging detected correctly all 54 biopsy-confirmed malignant breast lesions (down to 2 mm) among 207 BI-RADS 4 participants (NPV = 100%), while detecting “likely malignancy” in the biopsy region of 31 participants with negative biopsy (PPV = 63.5%), irrespective of breast density.

**Key messages:**

- *MUST detected all 54 biopsy-confirmed malignant lesions in 207 BI-RADS 4 patients (NPV=100%)*
- *MUST detected some malignancy in the region of biopsy in 31 cases with negative biopsy (PPV=63*.*5%)*
- *The diagnostic performance of MUST was independent of breast density*

## Introduction

Breast cancer is recognized as a major public health concern [1,2], leading the World Health Organization (WHO) to launch the Global Breast Cancer Initiative Framework in 2023, aiming to reduce breast cancer mortality by 2.5% annually and prevent 2.5 million breast cancer deaths globally by 2040 [3]. In this effort, the biggest challenge has been seen as the establishment of effective screening procedures that may detect breast cancer early and reduce mortality [4–6]. At present, Digital Mammography (DMG) is the dominant screening modality that is viewed as the “gold standard” for this purpose [6]. However, a notable drawback of DMG screening is “overdiagnosis”, which imposes unnecessary burdens on patients and healthcare systems [7–9]. Overdiagnosis is the detection of malignancies that would not have become clinically relevant in a patient’s lifetime, either because of their small size and the possibility of being eliminated by the immune response or because of having very slow growth to become threatening for metastasis. Estimating the rate of overdiagnosis is difficult due to methodological complexities, with reported estimated rates ranging from 12% to 40% [9,10], while the benefit-to-harm ratio is deemed favorable [11,12].

Among the diagnostic categories established by the Breast Imaging Reporting and Data System (BI-RADS) [13], the 4^th^ one is the most crucial for the issue of overdiagnosis, because it includes suspicious mammographic findings over a wide range of assessed probability of malignancy (2% - 95%) that cannot be fully characterized non-invasively [14]. The BI-RADS 4 category is further stratified into sub-categories: 4A (2 - 10%), 4B (10 - 50%) and 4C (50% - 95%) [15]. The overall malignancy risk in BI-RADS 4 patients is about 21% [16,17]. The BI-RADS 4 patients are referred to biopsy in standard clinical practice. The reported average statistics indicate that about three-fourths of biopsies are negative and can be potentially avoided if a suitable diagnostic technology can be found to serve as an adjunct to DMG to improve the specificity of the screening process. In addition to the low specificity, the sensitivity of DMG is low for dense breasts (about 40% of all cases), bringing the overall sensitivity of DMG to modest levels [18–20]. Some sensitivity improvements have been achieved with the introduction of 3D-DMG (tomosynthesis), but at the cost of lower specificity, while serious challenges remain for dense breasts [21–24].

To address the DMG limitations for dense breasts, echo-mode ultrasound (hand-held B-mode) has been used as an adjunct to DMG to assist in the diagnosis for challenging cases [25–30]. This has led to some diagnostic improvement in certain cases, although such improvement depends crucially on the skill/experience of the physician/operator. Breast MRI has also been actively explored for this purpose and has demonstrated significant improvements in sensitivity (more detections of malignancies) – although at the cost of lower specificity (more false-positive detections) [31–35]. In addition, MRI remains an expensive modality that is not very accessible to the general population and requires injection of contrast agent. Several other diagnostic imaging modalities have been explored for this purpose over the last 30 years (e.g. ultrasound-based elastography [36–39], semi-automated whole-breast ultrasound [40–42], mammography with contrast agent [43,44], molecular/gamma imaging [45], breast X-ray CT [46], optoacoustic imaging [47] and others), but none has been shown to offer significant reduction of DMG false-positives without compromising sensitivity.

This serious limitation of current modalities of breast cancer screening/diagnostic imaging has provided the motivation for the development and clinical evaluation of the novel diagnostic imaging technology of Multimodal UltraSonic Tomography (MUST) that seeks to improve significantly the specificity of DMG, without reducing the sensitivity, when used as an adjunct in the screening process [48–53]. This paper presents results of a recent clinical trial (#EUDAMED/CIV-ID CIV-GR-22-05-039513) that demonstrate the ability of the novel MUST imaging modality to *reduce significantly the number of DMG false-positives* (when used as an adjunct) in a cohort of BI-RADS 4 patients referred to biopsy. Note that the stereotactic biopsy results offer an excellent form of “ground truth” (in the region of biopsy) to evaluate any new or current breast diagnostic imaging modality.

## Methods & Materials

### Study design and enrollment of participants

The registered clinical trial EUDAMED/CIV-ID CIV-GR-22-05-039513 was carried out in accordance with the Code of Ethics of the Declaration of Helsinki and was approved by the Research Ethics Committee of the Hippocration Hospital of the University of Athens, School of Medicine. Informed consent was obtained from all participants (after enrollment screening using the specified inclusion and exclusion criteria), whose privacy rights are protected. Data from female participants were collected between March 7, 2023 and June 18, 2024 at the Breast Unit of the Department of Surgery in the Hippocration Hospital (“off-label” use for research purposes only). The sample size calculation was based on the width of the 95% confidence interval (CI) for the primary endpoint, assuming a 20% prevalence of malignant breast lesions and an expected MUST specificity of 75%, resulting in 199 participants required to achieve a 95% Clopper-Pearson CI for specificity with a width of 0.12. Assuming a 5% dropout rate, a total of 210 participants were planned to be enrolled in the study. Of those, 3 did not proceed to biopsy; thus data from 207 participants with average age of 52.86 (10.16) years were analyzed. All were White and the majority (58.6%) was postmenopausal. Breast densities varied within the cohort and covered all density grades (with the slight majority exhibiting dense breasts), as reported in the Results section. All participants were classified by radiologists as BI-RADS 4, based on their mammogram, and were referred to biopsy according to standard clinical practice. Prior to biopsy, they underwent MUST scanning. The participants were surveyed regarding comfort during MUST scanning.

### Ground-truth data (reference standard)

After the MUST scan, stereotactic biopsy was performed using the Mammotest Breast Biopsy System (Fischer Imaging Corporation) and tissue samples were retrieved under local anesthesia using the *Intact****™*** Breast Lesion Excision System (Intact Medical Corporation) with 20 mm basket to remove the mammographically detected lesion in the specimen. The histopathology of the biopsy specimens identified the malignant or benign lesions. The biopsy data constitute the “ground truth” for evaluating the MUST imaging results in the biopsy region.

### Brief description of the MUST imaging system

The MUST breast imaging technology has been developed and tested over the last 15 years, and it is akin to “*CT with transmission-mode ultrasound*”, as described in previous publications [18–24]. Briefly, MUST performs 3D tomographic scans of the pendant breast in a precisely controlled water-bath of constant temperature that is continuously de-ionized, degasified, filtered and sterilized. Transmitted ultrasound pulses of specially designed sequences traverse the breast tissue in the water-bath and are received by transducers on the opposite side for all azimuths over 20 cm (field of view) for each view-angle. This is repeated for multiple view-angles and elevations to scan the entire 3D volume of the breast in coronal planes. The separation between adjacent coronal planes is currently set at 4 mm (adjustable). The in-plane pixel size is 0.5 mm x 0.5 mm. The scan takes about 10 minutes for most breasts but twice as long for very large breasts. The MUST images are available within 1 min. All safety precautions are taken to eliminate electromechanical or hygienic risks for the patients. The MUST scan is comfortable, as it does not require compression or contact of the breast.

### Data analysis & image formation

Waveform changes in the received pulses are analyzed relative to their transmitted counterparts to extract the acoustic attributes of refractivity (based on measurements of speed of sound in tissue relative to water), and frequency-dependent attenuation at each tissue voxel (0.5 mm x 0.5 mm x 4 mm). The values of these acoustic attributes are calibrated relative to water-through transmission at fixed water temperature to retain global numerical validity for comparative diagnosis across different subjects, locations and times. The acoustic attributes (modes) of refractivity and frequency-dependent attenuation quantify the micromechanical properties of the cellular microstructure in each tissue voxel [41,42] and yield a set of “multimodal” images for each coronal plane of the breast. Fusion of these multimodal images, using a properly developed data-based algorithm, yields the “***Advisory Diagnostic Image” (ADI)*** for each coronal plane that depicts the “likelihood of malignancy” at each tissue voxel through a computed index that exceeds 1 when the “likelihood of malignancy” is deemed very high. The 3D stack of coronal ADIs allows fast and unambiguous examination of the entire breast volume and detection of suspected malignant lesions at 3D fixed-coordinates, independent of human operator or subjective interpretation (see Figure 1). The integration of this advisory information into the clinical workflow is the responsibility of the physicians, but it is expected to improve significantly the specificity of the screening process, without compromising the sensitivity, and shorten the workflow. The findings of the MUST ADIs in the biopsy region were evaluated against the “ground-truth” of the biopsy results (reference standard for this clinical trial). The correspondence of the biopsy location (marked on the mammogram) with the respective location on the 3D stack of MUST coronal images was assisted by a software tool developed for this purpose.

**Figure 1.**
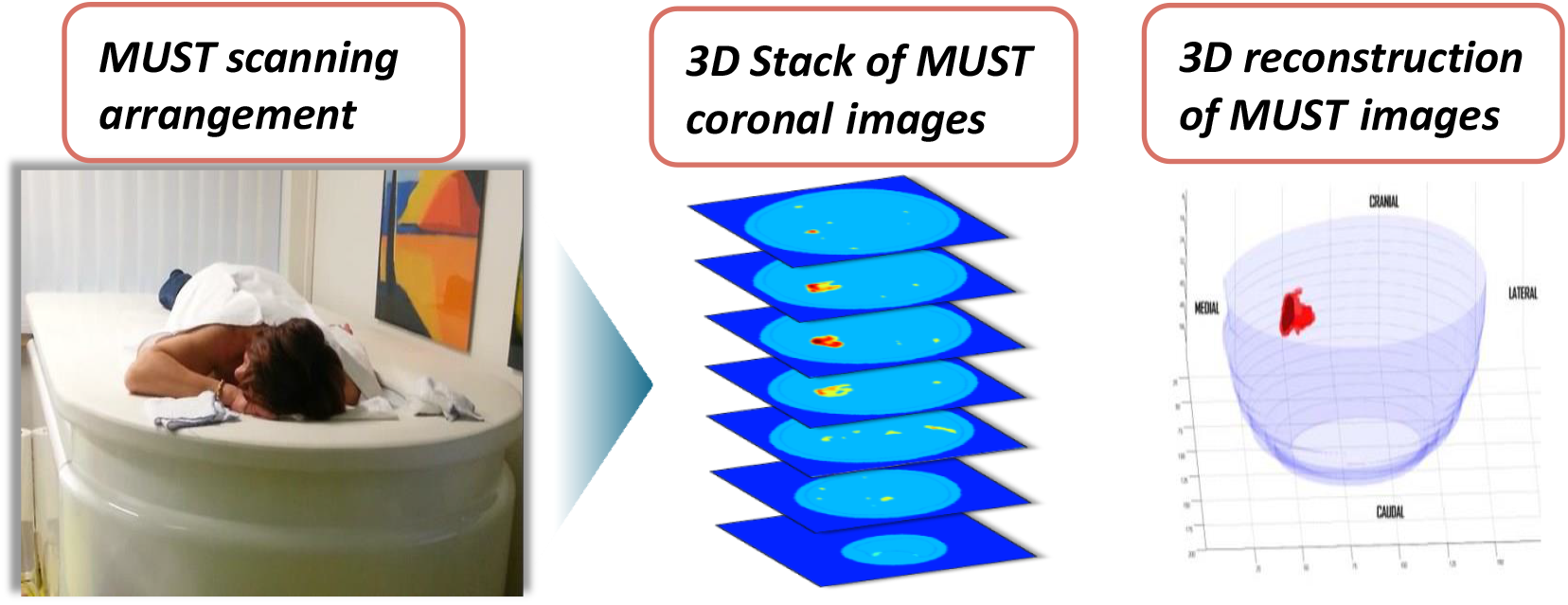
Illustrative schematic of MUST imaging process.

### Statistical analysis

The traditional diagnostic metrics of *sensitivity, specificity, Positive Predictive Value (PPV), Negative Predictive Value (NPV) and Accuracy* were used to quantify the benefits in diagnostic performance achieved by using MUST imaging as an adjunct to DMG. Furthermore, we computed the performance metrics of LR+/LR-defined as:

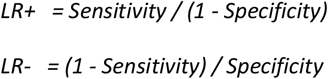

which are considered useful performance metrics [54,55] and are contained in the registered Clinical Investigation Report. To explore possible associations between breast density (using ACR scores 1-4) and biopsy outcome or MUST ADI outcome, the p-values of mean-difference t-tests were also computed.

## Results

Table 1 summarizes the biopsy results for the 207 participants that revealed 54 cases of malignant lesions (37 Ductal Carcinomas in Situ (DCIS), 15 Invasive Ductal Carcinomas, 1 Invasive Lobular Carcinoma and 1 Mucinous Carcinoma), along with the findings of the MUST ADIs in the biopsy region. The most important finding is that *all biopsy-confirmed malignant lesions were correctly identified by the MUST ADI as “likely malignant” (Negative Predictive Value (NPV) or sensitivity of 100%)*. There were 31 cases among the 153 negative biopsies where MUST ADI detected “likely malignancy” within 20-mm distance from the point of biopsy, yielding Positive Predictive Value (PPV) of 63.5% and specificity of 79.7%. The latter cases are viewed as “false-positives” for MUST imaging in this study, although the inherent inaccuracies of the biopsy process may also be noted.

**Table 1.**
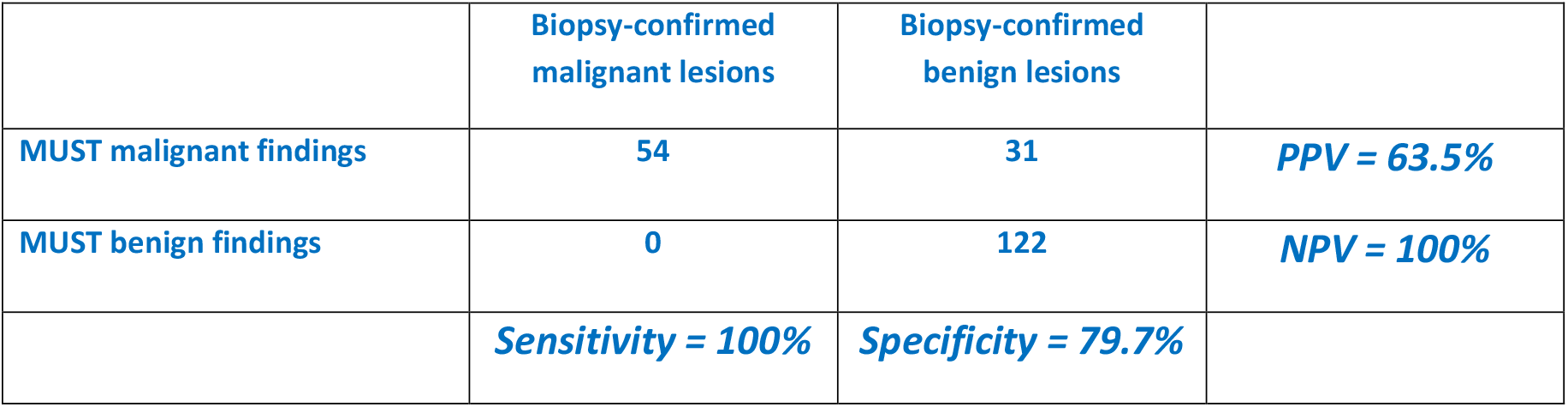
Biopsy results and MUST ADI findings.

We summarize below the main findings of this clinical study as reported in the official Clinical Investigation Report of the Clinical Trial registered under EUDAMED/CIV-ID CIV-GR-22-05-039513:

1. MUST demonstrated 100% sensitivity and NPV (95% CI: 99.96% - 100.00%).
2. MUST demonstrated specificity of 79.7% (95% CI: 78.94% - 80.52%) and PPV of 63.5% (95% CI: 62.58% - 64.47%). These compare with specificity and PPV of 26.1% for DMG.
3. MUST demonstrated 85% accuracy (95% CI: 84.31% - 85.71%), due to large reduction of false-positives.
4. The computed LR+ and LR-performance metrics were found to be 4.94 (95% CI: 3.68 - 7.02) and 0.00 (95% CI: 0.00 - 0.07), respectively, indicating that a negative MUST result effectively rules out malignancy, and a positive MUST result substantially increases the likelihood of malignancy.
5. The size of the MUST-detected malignant lesions reached down to 2 mm in maximum dimension.
6. There were 135 cases (65.2%) of dense breasts (106 of ACR 3 and 29 of ACR 4), indicating the efficacy of MUST for dense breasts as well.
7. No adverse events or device adverse events were reported, indicating the safety of MUST.
8. The majority of patients (56.2%) experienced no discomfort during the MUST procedure, while 34.8% reported slight discomfort, and only 9% reported moderate (7.6%) or substantial (0.9%), or severe discomfort (0.5%), indicating that the procedure was well-tolerated by the participants.

The main biopsy-identified 153 benign lesions were composed of the following types: 43 benign calcifications (20.5%), 30 apocrine metaplasia (14.3%), 27 fibroadenomas (12.9%), 14 flat epithelial atypia (6.7%), 12 ductal hyperplasia without atypia (5.7%), 7 atypical lobular hyperplasia (3.3%), 7 intraductal papillomas without atypia (3.3%), 5 atypical ductal hyperplasia (2.4%), 4 sclerosing adenosis (1.9%), 3 ductal ectasia (1.4%), 1 lipophagic granuloma (0.5%). Table 2 shows the primary benign lesions identified through biopsy for the 31 cases of MUST false-positive findings. About half were flat epithelial atypia (29.03%) and fibroadenomas (19.35%).

**Table 2.** Primary benign lesions identified through biopsy for the 31 cases of MUST false-positive findings.

| Biopsy-confirmed benign lesions | Number of MUST false-positives | Percentage of all benign lesions |
| --- | --- | --- |
| Flat Epithelial Atypia | 9 | 29.03% |
| Fibroadenoma | 6 | 19.35% |
| Apocrine Metaplasia | 5 | 16.13% |
| Benign Calcifications | 5 | 16.13% |
| Atypical Ductal Hyperplasia | 2 | 6.45% |
| Atypical Lobular Hyperplasia | 2 | 6.45% |
| Sclerosing Adenosis | 1 | 3.23% |
| Ductal Hyperplasia without Atypia | 1 | 3.23% |

To explore possible associations between breast density and biopsy outcome or MUST ADI outcome, the p-values of mean-difference two-sided t-tests (using the ACR density score in the scale 1-4) were computed for the following density comparisons: Positive vs Negative biopsy [mean (SD): 2.76 (0.73) vs 2.53 (0.75), p= 0.055], Positive vs Negative MUST ADI [mean (SD): 2.78 (0.72) vs 2.57 (0.76), p= 0.052], True-positive vs False-positive MUST ADI [mean (SD): 2.53 (0.75) vs 2.66 (0.78), p= 0.448]. Thus, no significant association with breast density was found in any of these group comparisons, although two of them were borderline at 95% significance level.

To address possible concerns about the suitability of the t-test for the ACR scores, categorical analysis was also performed using the Chi-square statistic, whereby ACR scores 1-2 or 3-4 were lumped together into “non-dense” or “dense” category, respectively. No statistical significance was found for these density comparisons as well.

An illustrative example of MUST imaging is presented in Figure 2, where the mammogram of the left breast of participant #151 (cranio-caudal (CC) and medio-lateral-oblique (MLO) views) are shown, along with 13 coronal ADI (4 mm apart). This participant presented clusters of micro-calcifications in the central area of zone B (area marked with orange circles in the mammogram) and had ACR density score 3 (i.e. heterogeneously dense). The marked area (biopsy region) corresponds to the central area of coronal ADI 5-8 that is also marked with orange circles, as determined by the software tool that we have developed for this purpose. The latter ADI area contains the two malignant lesions identified by histopathology: an Invasive Ductal Carcinoma (IDC) and a Ductal Carcinoma in Situ (DCIS). Several additional “likely malignant” detections are seen that are outside the region of biopsy and cannot be evaluated histopathologically. These exemplify the frequent finding of additional “likely malignant” detections away from the region of biopsy (>20 mm from putative point of biopsy).

**Figure 2.**
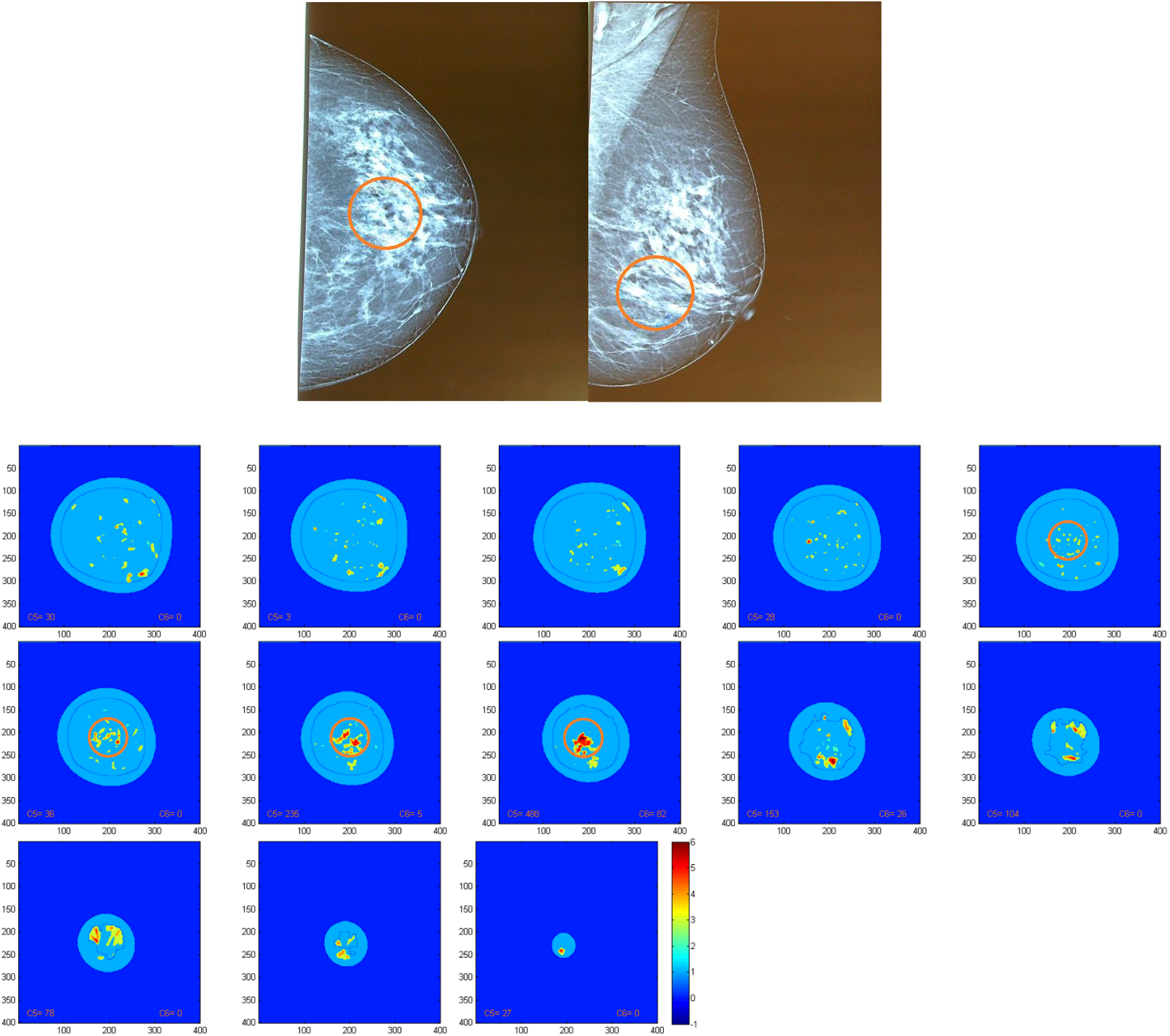
Top: the CC (left) and MLO (right) views of the mammogram of the left breast of participant #151 presenting micro-calcification clusters in the area marked with orange circles, where the biopsy identified two malignant lesions: an IDC and a DCIS. Bottom: 13 coronal MUST ADIs (4 mm apart) depicting the biopsy-confirmed IDC and DCIS lesions in red at the central region of ADIs 5-8 within the orange circles that demarcate the biopsy region (marked also on the mammogram). Some additional “likely malignant” lesions are detected by MUST ADI outside the region of biopsy (>20 mm from the putative point of biopsy).

Visualization of the MUST-detected *“likely malignant”* lesions within the breast volume may be facilitated by the sagittal display of the MUST ADIs, shown in Figure 3 (36 sagittal slices, 5-mm apart, displayed from medial-to-lateral side).

**Figure 3.**
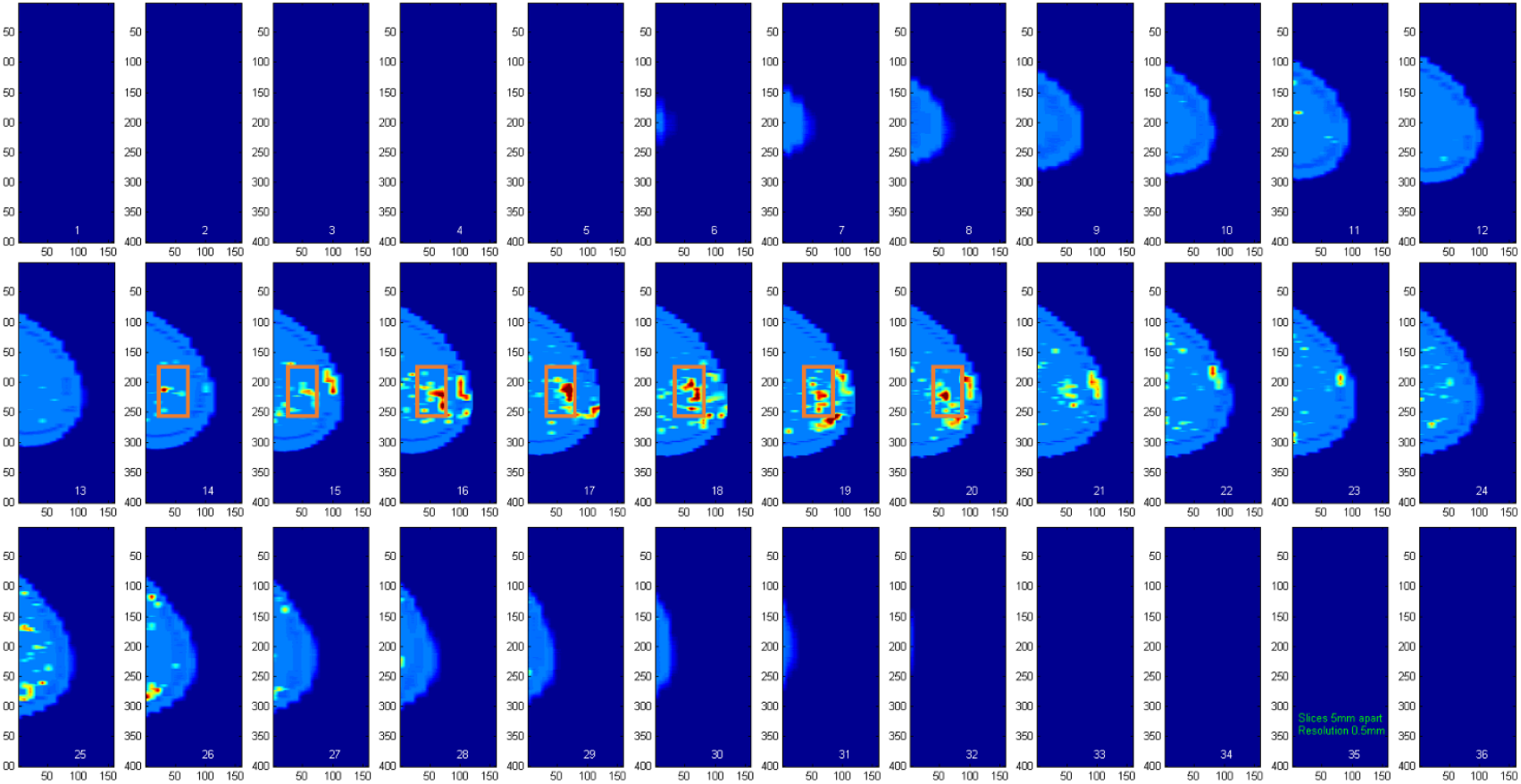
Sagittal display of the ADIs (36 slices displayed from medial-to-lateral side, 5-mm apart) for the left breast of participant #151 with biopsy-confirmed IDC and DCIS lesions seen on sagittal slices 14-20 within the orange rectangle. Additional “likely malignant” MUST detections are seen outside the biopsy region.

## Discussion

This paper presents the results of the first clinical trial (EUDAMED/CIV-ID CIV-GR-22-05-039513) evaluating the performance and safety of the MUST imaging device, used as an adjunct to DMG, in discriminating benign from malignant breast lesions identified through biopsy in a cohort of 207 BI-RADS 4 patients that provides adequate statistical power for the key findings, which are the following:

- ***MUST successfully characterized all biopsy-confirmed malignant lesions (100% sensitivity and NPV)***.
- ***MUST characterized most biopsy-confirmed benign lesions as non-malignant, yielding PPV = 63*.*5% and specificity of 79*.*7% (compared to DMG specificity and PPV of 26*.*1%)***.
- ***The overall diagnostic accuracy of MUST was 85%, due to large reduction of false-positives***.

Furthermore, the obtained diagnostic performance metric LR+ = 4.9 suggests that a positive MUST result substantially increases the likelihood of malignancy, while the obtained LR-= 0 confirms that a negative result effectively rules out malignancy. It is generally recognized that a low LR-is indicative of a highly reliable diagnostic test [54,55].

These results demonstrate that MUST, as an adjunct to DMG, can improve significantly the clinical management of BI-RADS 4 patients by reducing the number of biopsies by almost 80% (compared to DMG alone) without compromising sensitivity or NPV. By reducing unnecessary biopsies, MUST offers considerable benefits to the patients and reduces the clinical workflow and the economic burden on healthcare systems. In addition, the MUST procedure was well-tolerated by the participants, with 91% reporting no or slight discomfort (no breast compression is involved), while the absence of adverse events suggests that the device is safe for clinical use. Finally, we note that the MUST device is fully automated (no operator dependence or subjective interpretation), safely repeatable (no ionizing radiation) and requires no contrast enhancement. These results document an improved risk/benefit ratio for the MUST device when used as an adjunct to DMG.

The unique capability of MUST to detect and differentiate automatically cancerous lesions in the breast relies on the long-standing observation that malignant lesions exhibit different properties of sound propagation than benign lesions or normal tissues [56–59]. These differences in acoustic propagation properties can be quantified by speed-of-sound and frequency-dependent attenuation measurements. The latter can be extracted reliably by the MUST device using appropriate analysis of *waveform changes* in the received pulses traversing the breast tissue during the scanning process [48–53]. The extracted differences in these acoustic attributes (modes) are subsequently “fused” through a proprietary data-based algorithm to yield an index of “likelihood of malignancy” at each tissue voxel, leading to the generation of the MUST “Advisory Diagnostic Images” (ADI). This fusion algorithm was constructed using training and testing data from previous initial studies of BI-RADS 4 patients, some of which were published [48–53,60]. The results of the present study are consistent with our previously reported results from those smaller studies of BI-RADS 4 patients [52,53]. The aggregate published MUST results to date corroborate the exceptional sensitivity and improved specificity of MUST imaging as an adjunct to DMG. We note that the performance of MUST is not affected by breast density, as the results of the reported statistical testing suggest. We note that the information contained in the MUST images is fundamentally distinct from the one acquired through echo-mode ultrasound devices that have been used extensively in clinical practice to date, as the latter do not measure acoustic attenuation or refractivity – but only echogenicity.

An early transmission-mode ultrasound tomography system for breast cancer detection was developed at Mayo Clinic in the 70s and ‘80s, but its clinical testing was inconclusive [58,59]. Another system of 3D ultrasound tomography combined echo-mode ultrasound (distinct from MUST in its data collection and analysis procedures, thus lacking the automated lesion differentiation capability of MUST) has been described in the literature [61]. However, no significant clinical results have been reported yet on breast cancer detection by this system, and its use has been limited so far to automated whole-breast 3D imaging for breast density assessment and other breast volumetric measurements (without automated lesion differentiation capability).

We note that MUST also detected “likely malignancies” away from the region of biopsy, which represent a priority for investigation in the near future, as they will determine the clinical potential of MUST for breast cancer screening and may offer new insights into the important question of occult or indolent breast cancers. However, this task is very challenging, as biopsy cannot be ethically performed based on indications from a modality that is not yet certified for this purpose. Our current strategy is to examine with detailed histopathology *full mastectomies (ex vivo)* from volunteer patients who receive the surgery as part of the current standard of practice and also accept to receive the MUST scan prior to their surgery. The detailed histopathological examination of these mastectomies is a practically difficult task (especially for dense breasts) due to the practical limitations in the accuracy of the requisite “search and examination” procedure. Nonetheless, it may provide valuable “ground truth” for evaluating all MUST detections of “likely malignant” lesions, as well as allow performance comparisons with other breast imaging modalities.

## Conclusion

In a clinical trial involving 207 BI-RADS 4 participants (54 presenting malignant lesions) and using stereotactic biopsy results as “ground truth” (reference standard), MUST imaging as adjunct to DMG demonstrated much higher PPV (63.5%) than DMG alone (26.1%), while retaining perfect NPV (100%). Therefore, MUST as an adjunct to DMG may reduce unnecessary biopsies by approximately 80%, without compromising sensitivity or NPV.

## Data Availability

All data produced in the present study are available upon reasonable request to the authors

## Abbreviations

MUST: Multimodal UltraSonic Tomography,
BI-RADS: Breast Imaging Reporting and Data System,
MRI: Magnetic Resonance Imaging,
CC: cranio-caudal,
MLO: medio-lateral oblique,
ACR: American College of Radiology,
NPV: Negative Predictive Value,
PPV: Positive Predictive Value.

## Acknowledgements

*This work was supported by MUST Imaging Inc. and its subsidiary Mastoscopia S.A. which have built the MUST clinical prototype and supported this clinical trial. The authors thankfully acknowledge the valuable support offered by the staff of the Breast Unit of the Department of Surgery at the University of Athens, School of Medicine*.

